# Using Artificial Intelligence to Assess Treatment-Effect Heterogeneity in Pragmatic Cardiovascular Trials: Insights from TRANSFORM-HF

**DOI:** 10.64898/2026.01.16.26344310

**Authors:** Guangyu Tong, Changjun Li, Fan Li, Yukang Zeng, Stephen J. Greene, Kevin J. Anstrom, Jeffrey Testani, Robert J. Mentz, Eric J. Velazquez

## Abstract

**Background and Aims:** Pragmatic clinical trials are designed to assess interventions in real-world settings, and their broad inclusion criteria and clinical variability create valuable opportunities for exploring heterogeneity of treatment effects. In this context, TRANSFORM-HF was a pragmatic trial that found no overall survival difference between torsemide and furosemide in patients hospitalized with heart failure (HF), highlighting the importance of analytic strategies that can uncover clinically meaningful variation in treatment response.

The aim of this study was to evaluate whether baseline patient characteristics modify the relative survival benefit of torsemide versus furosemide using machine learning methods.

**Methods:** This was a post hoc analysis of the pragmatic, multicenter, open-label, randomized TRANSFORM-HF trial, which enrolled 2,859 patients hospitalized with HF across 60 US hospitals and randomized them to torsemide or furosemide. More than 50 baseline covariates were incorporated into a Bayesian Accelerated Failure Time model with Bayesian Additive Regression Trees (AFT-BART) to estimate individualized survival treatment effects (ISTEs). The outcome of interest was all-cause mortality over follow-up. Machine learning analyses estimated ISTEs for each patient and identified key baseline covariates modifying relative treatment effects. An exploratory decision tree was used to aid interpretability and to highlight influential effect modifiers.

**Results:** While the overall trial showed no survival difference, machine learning analyses revealed heterogeneity in treatment effects. Atrial fibrillation (AF), BNP/NT-proBNP levels, and prior loop diuretic use emerged as the most influential baseline modifiers. Patients without AF, with lower BNP levels, and without prior loop use showed suggestive benefit from torsemide (fitted mean ISTE up to 0.49; 95% CrI, –0.07 to 1.04), whereas patients with AF, elevated BNP and prior loop use showed relative benefit with furosemide (fitted mean ISTE –0.40; 95% CrI, –0.78 to –0.05). Although most subgroup effect estimates were statistically inconclusive due to wide credible intervals, split points were statistically significant, highlighting the importance of candidate modifiers.

**Conclusions:** In this exploratory AI-driven analysis of TRANSFORM-HF, we identified meaningful heterogeneity in diuretic response, with atrial fibrillation, natriuretic peptide levels, and prior diuretic use modifying relative survival benefit. These findings illustrate how integrating artificial intelligence into the analysis of pragmatic trials can move beyond average treatment effects to generate individualized, hypothesis-generating insights, and underscore the value of anticipating and systematically evaluating treatment-effect heterogeneity when designing and interpreting pragmatic cardiovascular trials.

**What is the clinical question being addressed?:** To apply artificial intelligence to detect treatment effect heterogeneity in the pragmatic TRANSFORM HF trial and to identify patient subgroups with differential benefit from torsemide versus furosemide, supporting precision medicine in heart failure.

**What is the main finding?:** Machine learning analyses within this pragmatic trial revealed substantial heterogeneity in the survival effects of torsemide versus furosemide. Torsemide appeared more beneficial among patients without atrial fibrillation and with lower BNP/NT-proBNP levels, whereas furosemide was favored in those with atrial fibrillation, higher BNP/NT-proBNP concentrations, and prior loop diuretic use. Exploratory analyses demonstrated that applying AI-driven methods to evaluate treatment effect heterogeneity within a pragmatic trial framework can leverage the inherent clinical diversity of pragmatic designs and potentially generate personalized and practice-relevant insights from pragmatic clinical research.

## Introduction

Pragmatic randomized trials are designed to evaluate the real-world effectiveness of interventions in in routine clinical practice, typically using broad eligibility, flexible delivery, and outcomes of direct relevance to patients and health systems. ^1–3^ They are increasingly used across cardiovascular care, and their inclusive designs naturally produce substantial clinical variation, center-level differences, and diverse patterns of adherence and co-interventions. As a result, heterogeneity of treatment effects (HTE) is often more pronounced and more clinically informative than in tightly controlled efficacy trials. ^4^ Rather than focusing only on a single average treatment effect (ATE), pragmatic trials can be planned and analyzed to characterize variation in treatment effect in routine practice.^3,4^

Traditional regression-based approaches are limited in estimating HTE because they require strong assumptions about model form, often test only a handful of pre-specified interactions, and are limited in their ability to capture high-dimensional, nonlinear relationships.^5,6^ In contrast, recent advances in machine learning, including modern AI techniques, provide a flexible and data-adaptive alternative that can incorporate many covariates, accommodate nonlinear relationships, and estimate individualized treatment effects with minimal overfitting.^7,8^ Although such analyses are exploratory and hypothesis-generating rather than definitive, they can yield insights that echo mechanistic research, generate testable hypotheses, and help design future confirmatory studies. They also allow neutral overall results from pragmatic trials to become opportunities for understanding treatment variation.

The Torsemide Comparison with Furosemide for Management of Heart Failure (TRANSFORM-HF) trial illustrates this potential.^9^ The trial was designed to address uncertainty regarding the comparative effectiveness of furosemide and torsemide, two loop diuretics that are central to symptom management in heart failure. TRANSFORM-HF enrolled 2,859 patients across 60 United States hospitals between 2018 and 2022. The trial population was clinically diverse, with a median age of 65 years, 36.9 percent women, and 33.9 percent Black participants. The primary analysis found no difference in 30-month all-cause mortality between torsemide and furosemide (HR 1.02; 95% CI 0.89–1.18).^9^ Similar findings were observed in subgroup analyses across different heart pre-specified subgroup analyses, including de novo vs. worsening chronic HF^10^ and ejection fraction categories.^11^

Despite these consistent results, the inherent heterogeneity of pragmatic design in TRANSFORM HF leaves open the question of whether all patients derive comparable benefit from either diuretic, or whether meaningful differences exist for specific subgroups. Indeed, existing trials and observational studies have produced mixed results. Some investigations have reported pharmacokinetic advantages of torsemide, including higher bioavailability, a longer half-life, and potential antifibrotic effects,^12^ while several meta-analyses and observational studies have suggested improvements in functional status, reductions in heart failure hospitalizations, and lower cardiac-specific mortality compared with furosemide.^13,14^ Other studies, particularly those using larger adjusted cohorts, have found no difference in all-cause mortality after accounting for confounding. ^15,16^ These mixed findings, together with the neutral overall result of TRANSFORM-HF, underscore continued interest in whether particular patient subgroups may respond differently to torsemide versus furosemide.

In this study, we apply machine learning–based HTE methods to TRANSFORM-HF to assess whether baseline clinical characteristics modify the comparative effectiveness of torsemide and furosemide. The goal is to characterize treatment effect variation using observed clinical features and to generate evidence that may inform more individualized approaches to diuretic therapy in heart failure. In doing so, we present a structured workflow for conducting and reporting HTE analyses in pragmatic cardiovascular trials.

## Methods

We analyzed the TRANSFORM-HF dataset and identified more than 50 baseline characteristics with clear definitions and less than 25 percent missingness for inclusion as covariates in the machine learning models. These variables include demographic factors (e.g., age, sex, race), clinical measures (e.g., systolic and diastolic blood pressure, heart rate, and BMI), laboratory values (e.g., sodium, potassium, creatinine, blood urea nitrogen, hemoglobin, and glomerular filtration rate) and common HF measures (e.g., LVEF, KCCQ, NYHA class, duration of heart failure), comorbidities (e.g., diabetes, atrial fibrillation/flutter (AF), hypertension, chronic kidney disease, chronic lung disease, myocardial infarction, coronary artery disease, peripheral artery disease, and stroke), and medication use (e.g., beta-blockers, ACE inhibitors, ARNi, or ARBs, MRAs, prior use of loop diuretics, as well as anticoagulants, antiplatelet therapies, and other treatments including insulin, GLP-1 agonists, SGLT2 inhibitors, hydralazine, digoxin, and long-acting nitrates), length of stay of index hospitalization and prior HF hospitalizations, the use of cardiac devices (e.g., defibrillators or cardiac resynchronization therapy), and socioeconomic and lifestyle factors were incorporated (insurance type, smoking status, and hospital type: teaching hospital or VA hospital).

Baseline characteristics across treatment groups were compared using the following tests: two-sample t-tests for continuous variables; Pearson’s chi-square for binary variables; joint Pearson’s chi-square for multi-level nominal variables; and Cochran–Armitage trend tests for ordinal variables. We then employed a Bayesian Additive Regression Tree (BART) model with nonparametric error term (see ^7^) within an Accelerated Failure Time (AFT) framework (AFT-BART-NP) to estimate individual survival treatment effects (ISTEs) of torsemide versus furosemide on 30-month all-cause mortality,^17^ using screened baseline covariates (including missingness indicators). The ISTE for a patient is defined as the difference between mean log survival times under torsemide and furosemide given baseline characteristics. The AFT-BART models the distribution of the log survival time, with the mean log survival time specified using an ensemble of regression trees.^18^ BART accommodates nonlinearities and interactions,^19^ with Markov Chain Monte Carlo (MCMC) sampling estimating individualized treatment effects as rate ratios in survival time. Specifically, we separately fit AFT-BART models in the torsemide and furosemide cohorts, used each model to predict an individual’s expected log survival time under both treatments based on their baseline covariates, and then estimate the ISTE as the difference between those two predicted log-survival times. To quantify ISTE uncertainty, we generated 1,000 posterior MCMC samples per individual from the AFT-BART model and reported 95% credible intervals. We compared the baseline characteristics between the 10% patients with the highest ISTE (favoring torsemide) and the 10% patients with the lowest ISTE (favoring furosemide). We then implemented an exploratory fit-the-fit approach through the classification and regression tree (CART),^8^ where the estimated ISTE was modeled as a response variable and patient characteristics most likely to influence this treatment difference were selected and used to categorize patients into different subgroups.

All analyses were performed on R 4.2.2. To aid interpretation of our exploratory classification and regression tree (CART) analysis, we developed an interactive Shiny App as a visual supplement to the findings (https://changjun.shinyapps.io/transformhfdecisiontool/) that allows users to input patient-level covariates and visualize the relative benefit for survival. This tool is intended solely for exploratory and illustrative purposes, and should not be applied to guide clinical decision-making until further validation is performed.

## Results

Among all baseline covariates included in the AFT-BART model, baseline characteristics were generally well balanced between groups (**Table 1**). The mean age was 64 years, and just over one-third of participants were women, with a slightly higher proportion in the furosemide group (39.0% vs. 34.8%, p = 0.022). Racial distribution was similar, with about one-third identifying as Black and over half as White, representing a diverse patient population. Clinical measures—including blood pressure, heart rate, renal function, electrolytes, and hemoglobin—were comparable, as were comorbidities such as diabetes (48%), atrial fibrillation/flutter (45%), chronic kidney disease (35%), and chronic lung disease (24%). Use of guideline-directed medical therapy was also balanced, with high rates of beta-blocker (79%), renin–angiotensin system inhibitors (60%), and mineralocorticoid receptor antagonist (36%) therapy. Most patients had reduced ejection fraction (≤40%, ∼70%) and were treated with guideline-directed therapies at similar rates. Notable differences included higher baseline KCCQ clinical summary scores among torsemide-treated patients (44.2 vs. 42.2, p = 0.021) and a greater history of prior CABG (17.3% vs. 13.9%, p = 0.014).

**Table 1.**
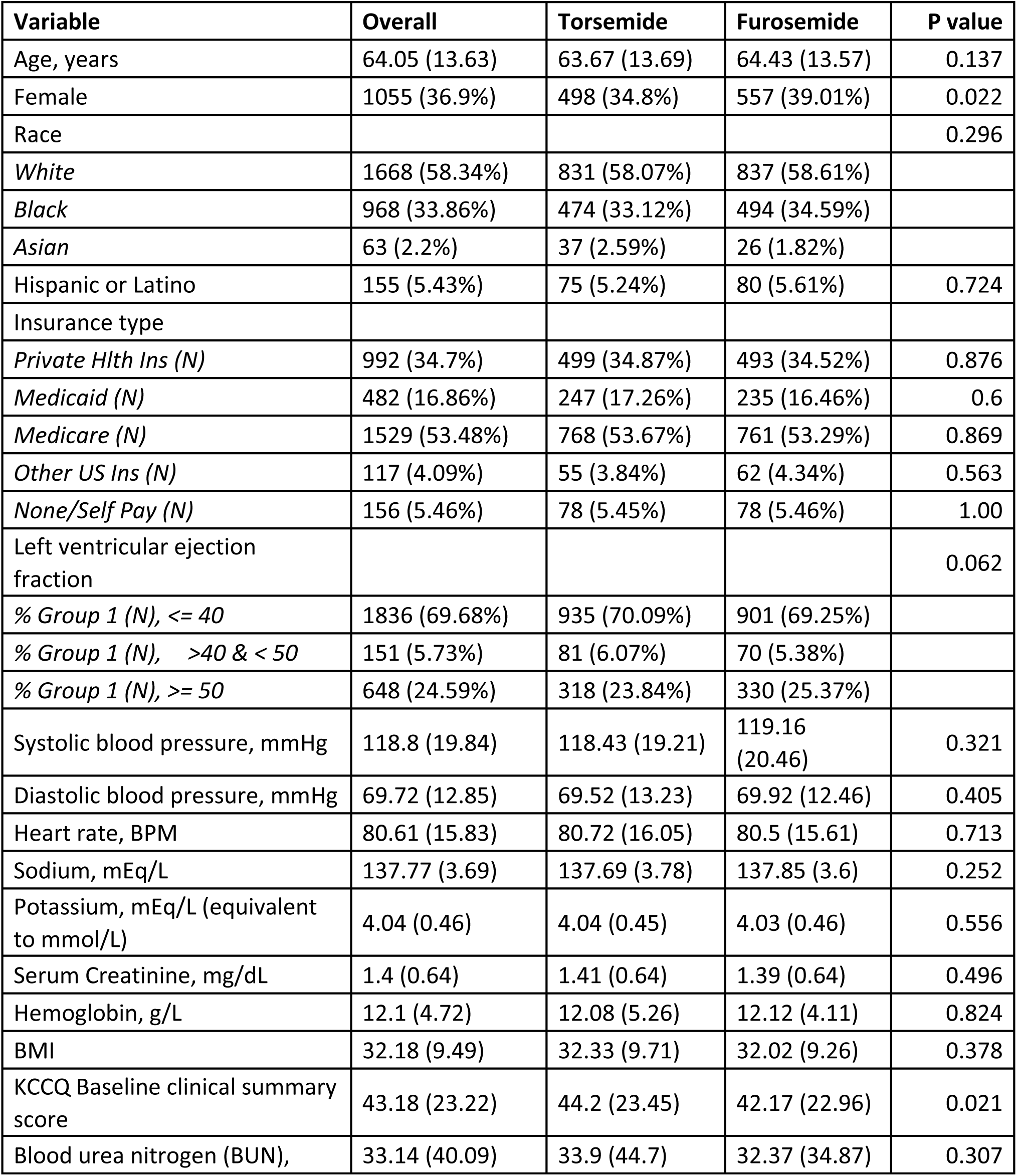
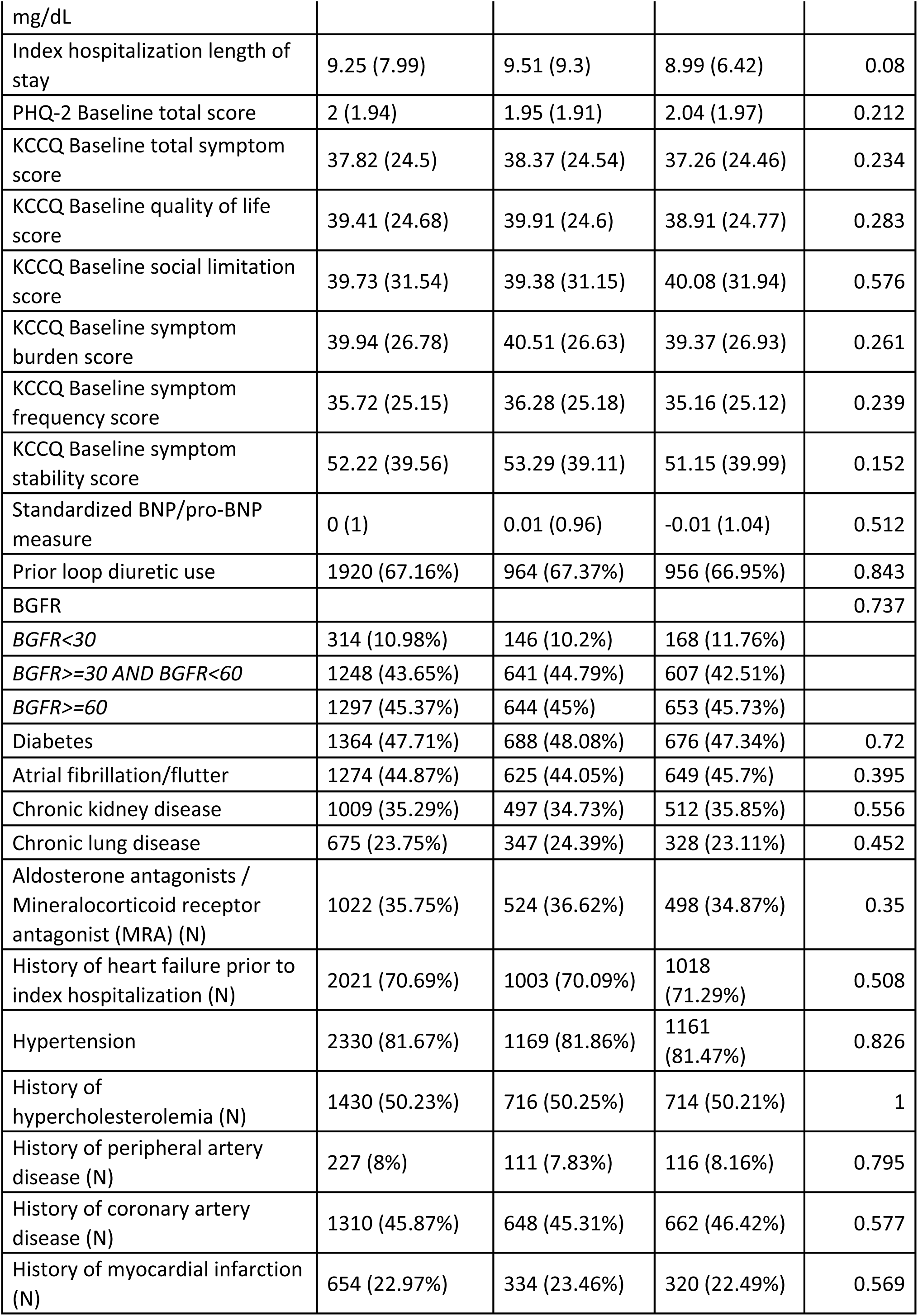
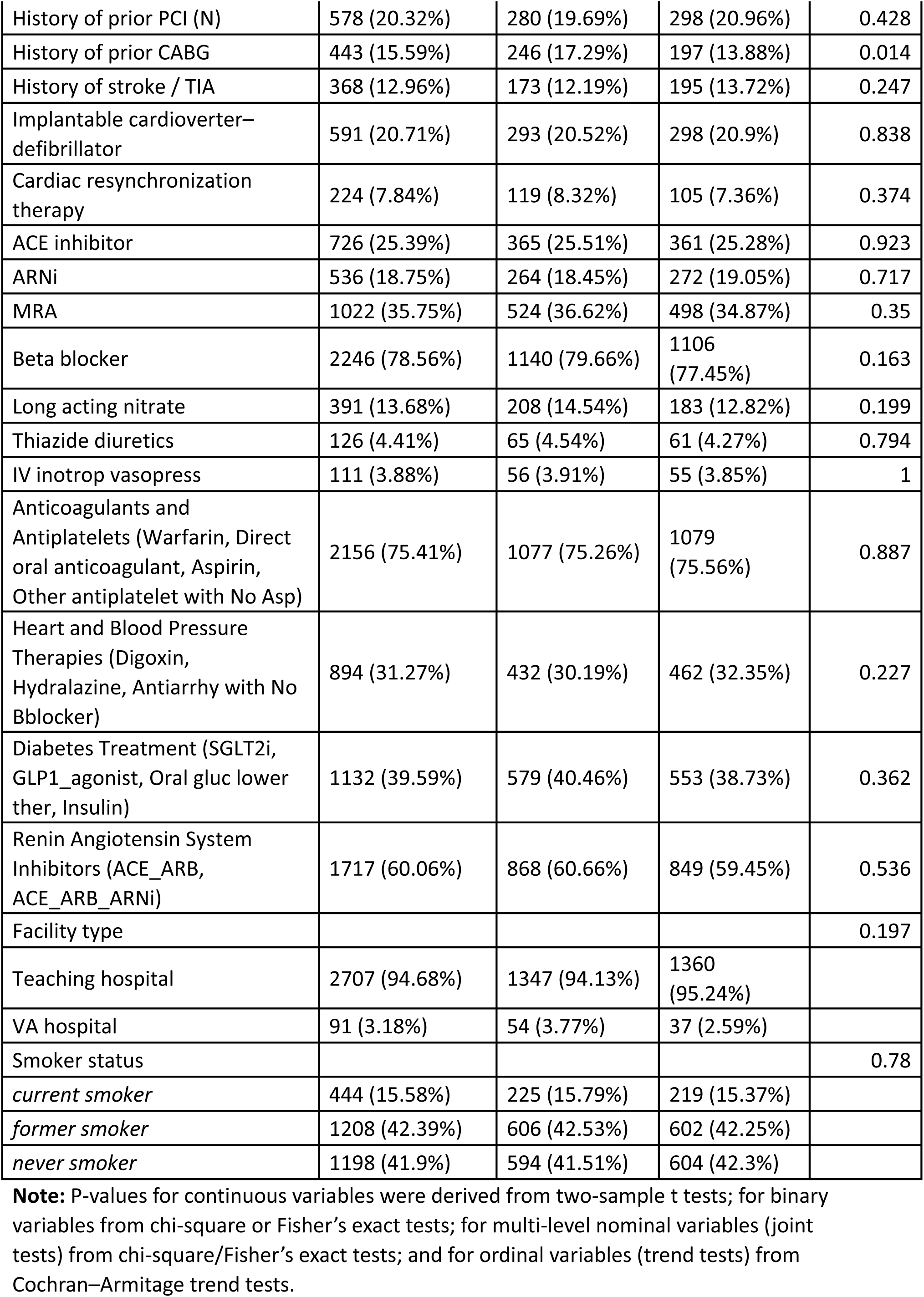
Descriptive statistics of baseline covariates of TRANSFORM HF.

By 30 months, among 1,431 patients randomized to torsemide, 373 died; among 1,428 patients randomized to furosemide, 374 died. **Figure 1** illustrates the estimated individual-level treatment effects (ISTE) on the log-survival time scale, obtained from our AFT-BART model. In panel (a), each bar in the histogram shows the fraction of patients with a given estimated effect, and the overlaid curve gives a smoothed density. The vertical dashed line at zero denotes no difference between torsemide and furosemide; estimates to the left of zero (negative values) indicate longer predicted survival under furosemide, whereas values to the right (positive) indicate a benefit for torsemide. The distribution is roughly symmetric around a mean ISTE of -0.02 log-days (95% CrI: [-0.34, 0.25]), indicating no appreciable difference in estimated survival benefit between furosemide and torsemide. Panel (b) depicts the sorted ISTEs where the solid blue dots represent the mean ISTE for each patient, and the shaded area indicates the 95% CrIs. On the x-axis, each index corresponds to one patient, where the leftmost points are those with the most predicted benefit from furosemide (negative ISTE), and the rightmost points are those with the most predicted benefit from torsemide (positive ISTE). Although a very small number of patients correspond to 95% CrIs that exclude the null, the dispersion of the ISTE estimates a non-trivial amount of treatment effect heterogeneity observed among the trial population.

**Figure 1.**
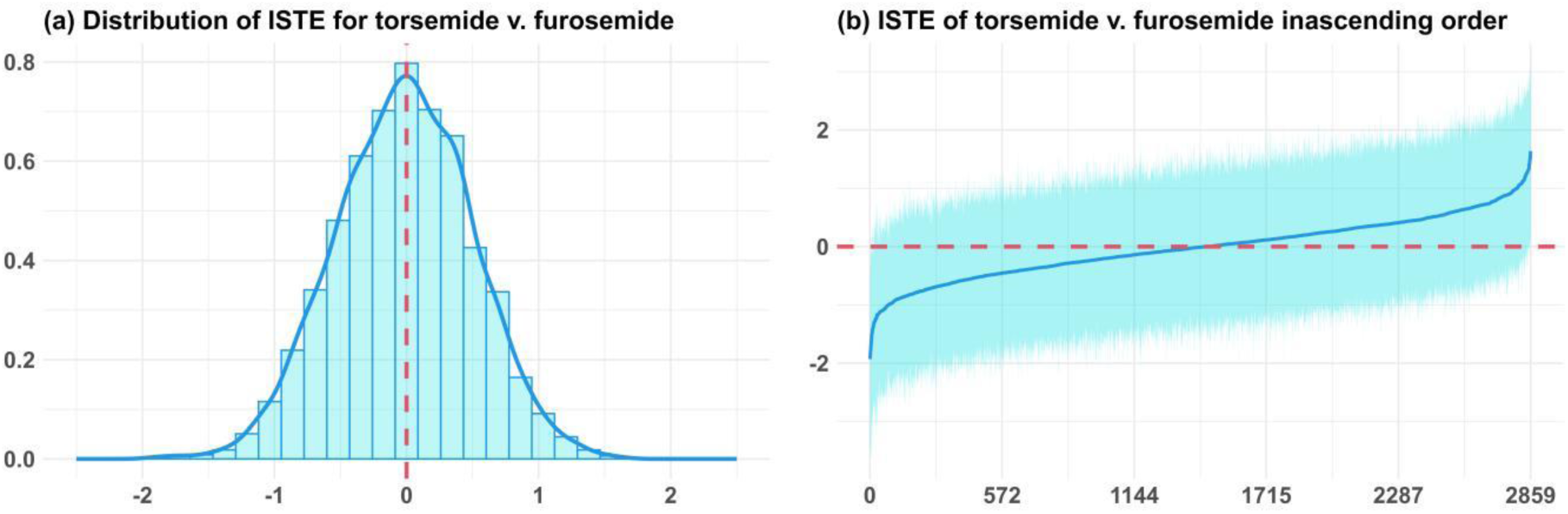
Distribution of ISTEs with credible intervals. Individual survival treatment effect (ISTE) of torsemide v. furosemide in the log time scale based on the AFT-BART model, with negative values favoring furosemide and positive values favoring torsemide. **Panel (a)**: histogram with density curve for the conditional average treatment effect (CATE) of torsemide v. furosemide. CATE represents ISTE=Expected log survival time under torsemide−Expected log survival time under furosemide. **Panel (b):** x-axis is the patient index, from 1 to 2,859, and y-axis is the CATE in the log time scale.

**Table 2** shows that, compared with patients in the bottom 10% predicted to benefit from furosemide, those in the top 10% predicted to benefit from torsemide are younger (mean 61.8 vs 66.1 years), more often Black and less often White or Hispanic, and present with lower standardized BNP, higher BMI, fewer prior loop-diuretic exposures, and shorter index hospital stays—findings that together suggest earlier-stage heart failure. They also exhibit fewer markers of advanced structural heart disease—less reduced EF (≤40%), lower prevalence of atrial fibrillation, markedly fewer prior ischemic events (myocardial infarction, PCI, CABG) and stroke/TIA, and less use of device therapies (ICD/CRT). Background pharmacotherapy patterns differ: ARNi, β-blockers, and renin–angiotensin system inhibitors are less common in the torsemide-benefit group, whereas MRA use is more frequent; PHQ-2 is modestly higher, with higher KCCQ symptom-stability but lower quality-of-life scores.

**Table 2.**
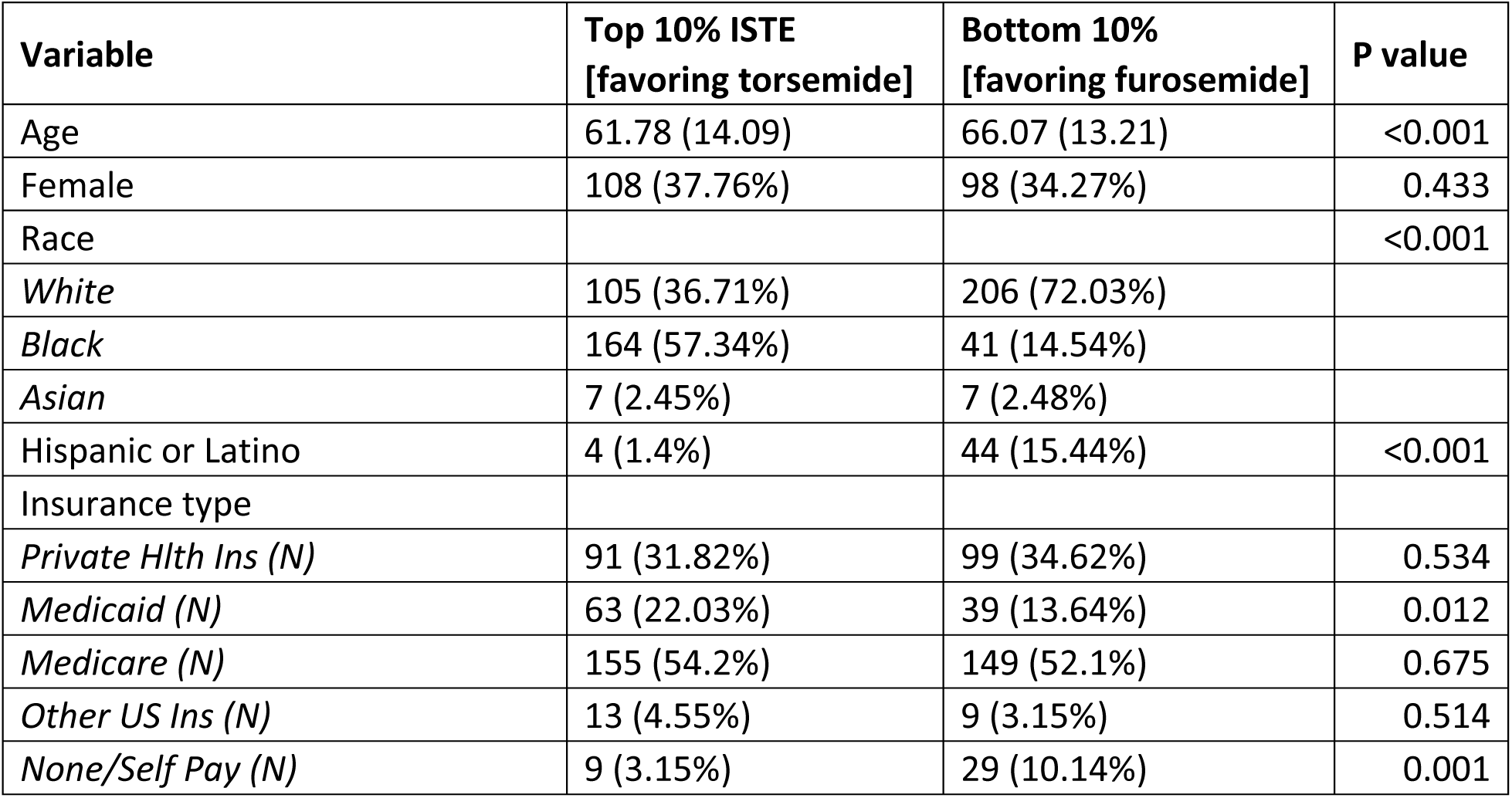
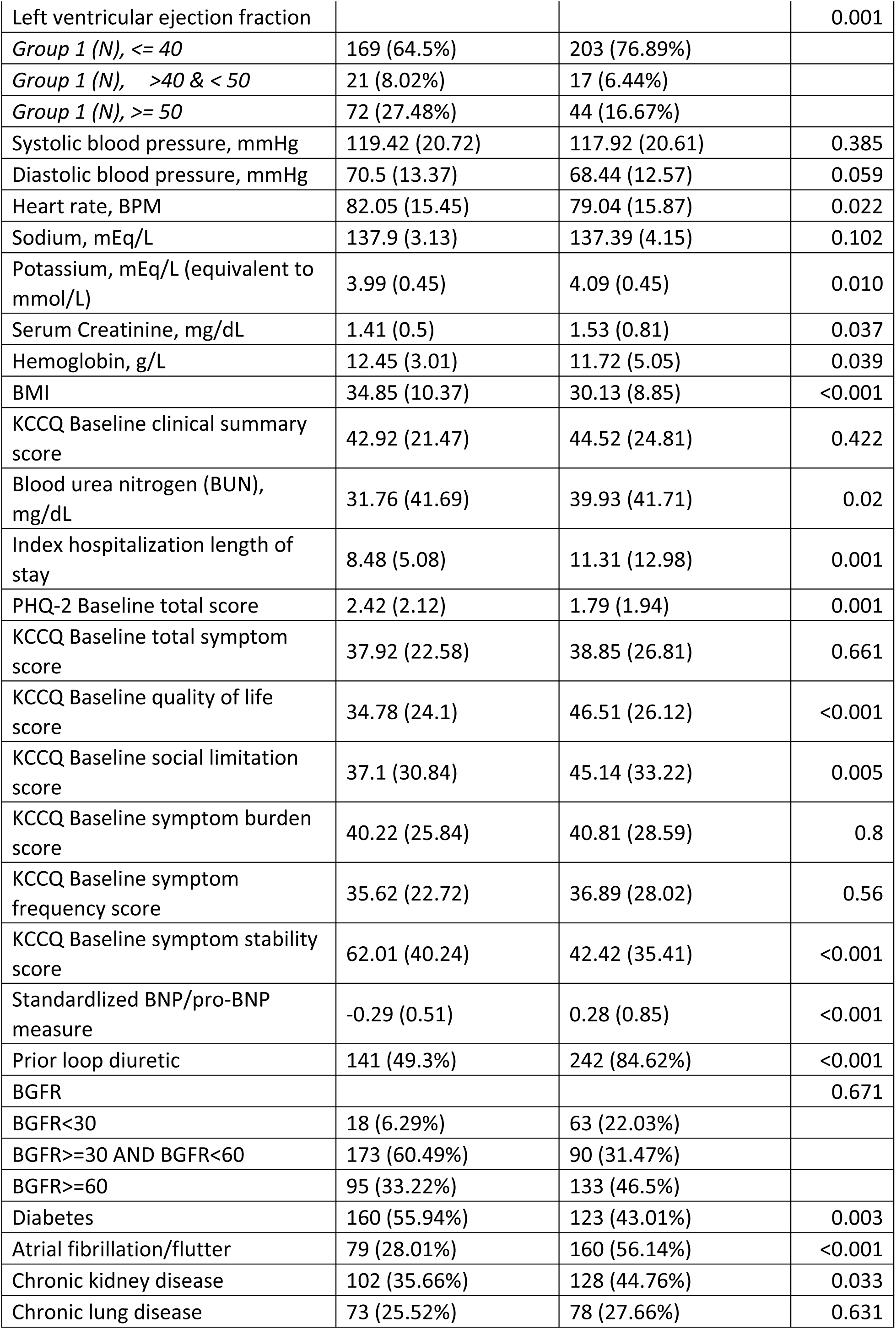
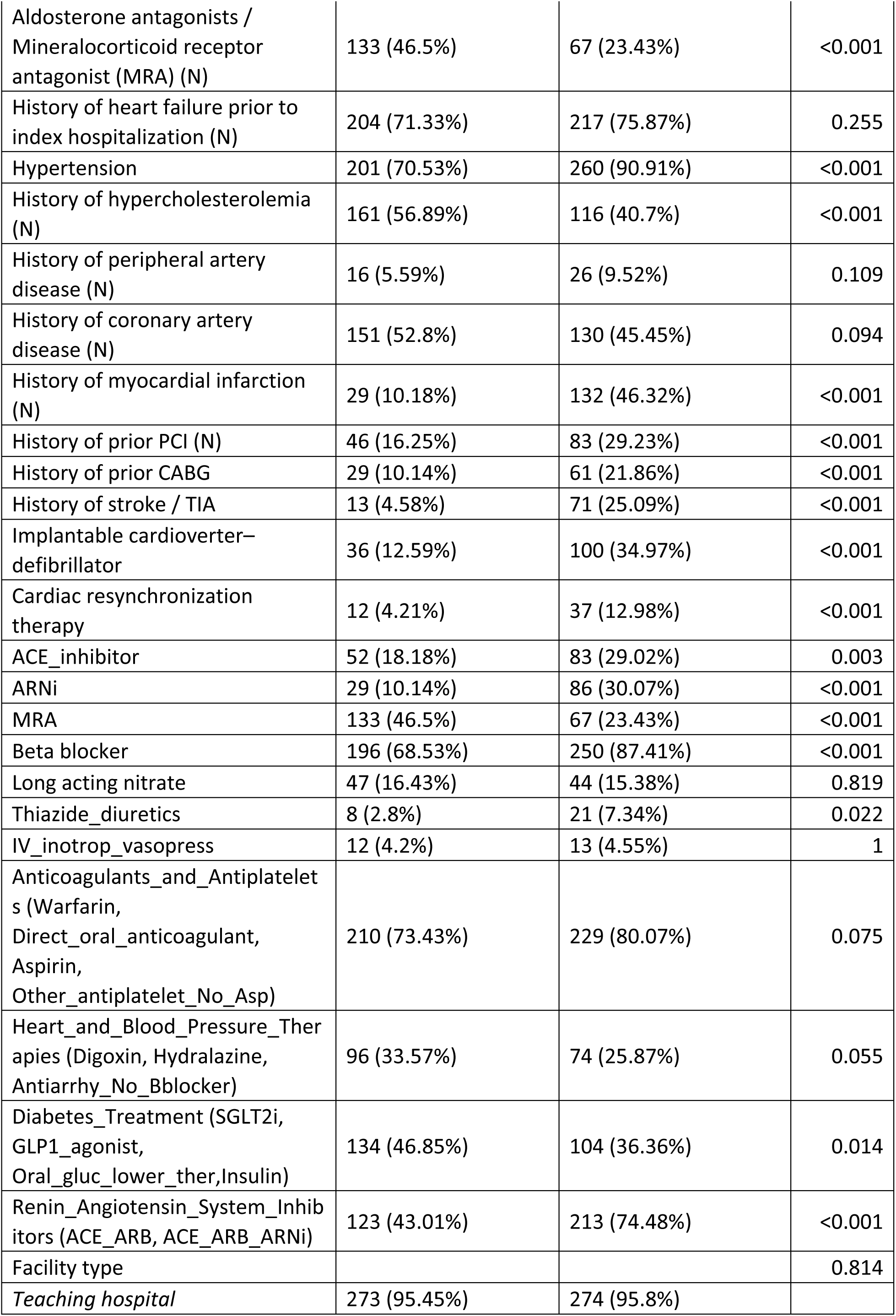
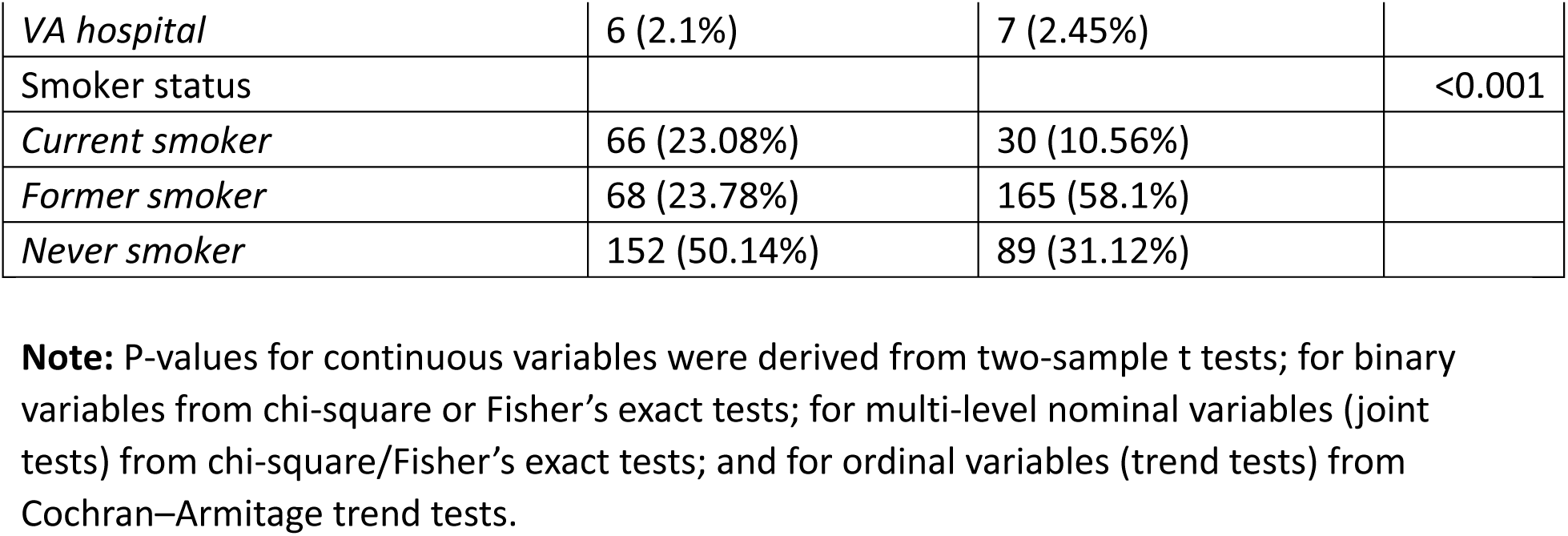
Comparison of baseline covariates among patients with the highest 10% IST Es (favoring torsemide) and the lowest 10% ISTEs (favoring furosemide).

Conversely, the furosemide-benefit group is older, more often White or Hispanic, shows higher congestion (BNP), greater burdens of AF, hypertension, ischemic and cerebrovascular disease, more device therapy and guideline-directed agents and longer hospital stays, alongside distinct insurance patterns indicative of socioeconomic disparities. Overall, these results suggest torsemide may yield the greatest incremental advantage in patients with relatively preserved health and less prior decongestive therapy, whereas furosemide may better serve those with a higher overall disease burden and entrenched guideline-directed treatments.

**Figure 2** presents a three-layer decision tree illustrating the most influential baseline patient characteristics associated with heterogeneity in individualized survival treatment effects (ISTEs) comparing torsemide versus furosemide. The root node splits patients based on a history of AF (p < 0.01). Among patients without AF (fitted mean = –0.23; 95% CrI: –0.54, 0.08; n = 1274), the next most influential factor was standardized BNP/NT-proBNP (split at – 0.25; p < 0.001). Those with higher BNP levels demonstrated more negative ISTEs (fitted mean = –0.40; 95% CrI: –0.78, –0.05; n = 498), suggesting a relative survival advantage with furosemide in this subgroup, while those with lower BNP showed smaller and more uncertain effects. Among patients with AF (fitted mean = 0.14; 95% CrI: –0.23, 0.50; n = 1585), BNP was again the most important modifier (p < 0.001). Within this branch, prior loop diuretic use further stratified responses (p < 0.01), with non-users showing suggestive benefits from torsemide (e.g., fitted mean = 0.49; 95% CrI: –0.07, 1.04; n = 200), whereas prior users had more attenuated effects. Although most specific subgroup effects were not statistically significant—likely reflecting smaller sample sizes in terminal nodes— the split points themselves (AF history, BNP levels, prior loop diuretic use) were highly significant, supporting their role as key baseline modifiers of differential diuretic response. These findings underscore the potential of data-driven methods to reveal heterogeneity that may guide future confirmatory analyses.

**Figure 2.**
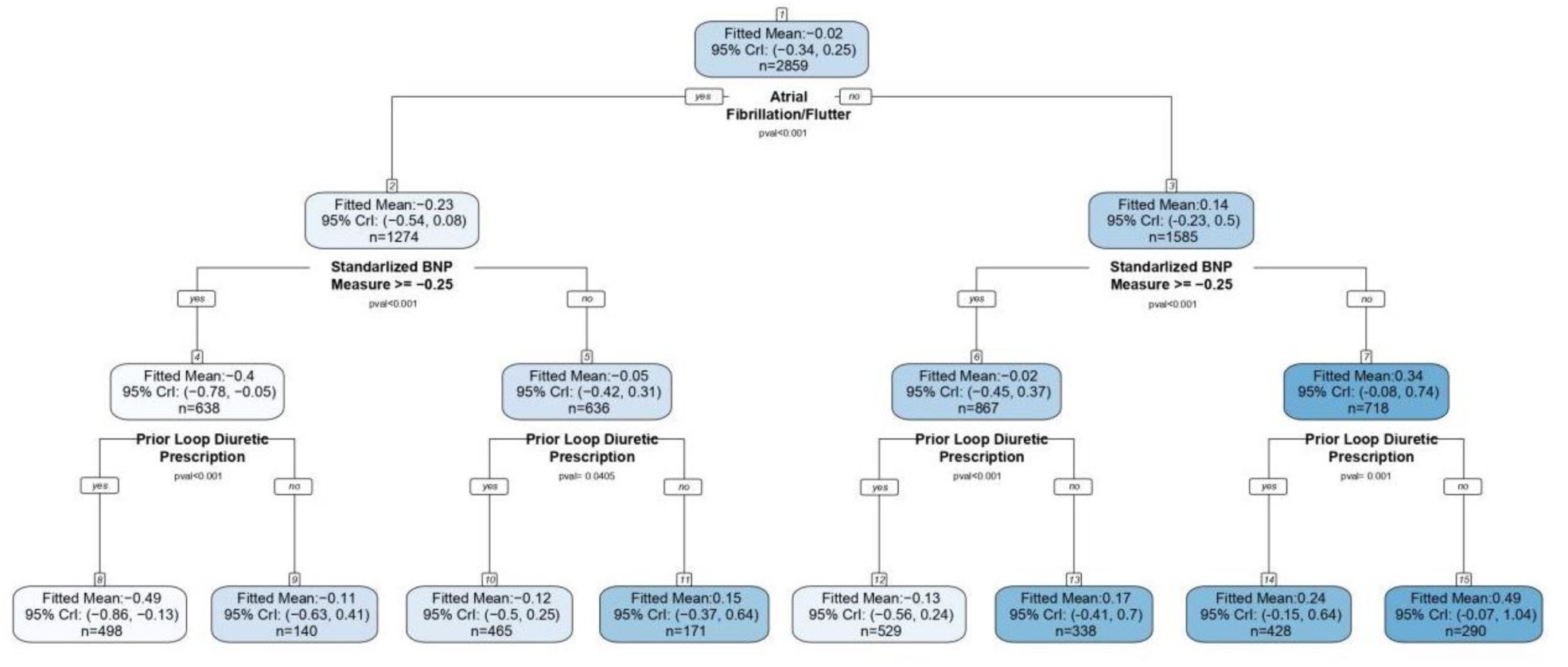
Classification and Regression Tree (CART) on ISTE for all-cause mortality at 30 months. CART model fit to the posterior CATE estimates (defined as the difference in log time to acute kidney injury). Values (fitted mean) in each node represent the posterior mean of the CATEs of the subgroup of patients described in that node; n in each node is the number of patients in that subgroup. The conditions below the nodes are rules used to divide subgroups, where “yes” indicates the condition is true, and “no” indicates the condition is false. Standardized BNP is the combined score of BNP and NTproBNP, where the value of -0.25 corresponds to BNP=751 or NT-proBNP=4,599. Deeper blue color indicates point estimates more favorable of torsemide, while lighter blue color indicates point estimates more favorable of furosemide.

**Figure 3** presents the subgroup-specific survival curves over 30 months for torsemide (teal) versus furosemide (pink), estimated from our fitted AFT-BART models. In each of the eight panels—corresponding to patient subgroups defined by individual survival treatment effects (identified in Figure 2)—the solid lines trace the posterior mean survival probability, and the shaded bands denote 95% credible intervals. While torsemide’s curve lies modestly above furosemide’s in subgroups 1, 2, 3 and 5 (and slightly below in subgroup 8), the two curves remain closely intertwined and their credible bands overlap almost entirely across all subgroups. Thus, despite visual hints of heterogeneity, there is no robust or statistically credible separation in survival probability between treatments in any subgroup. These graphical findings reinforce our CHR results and suggest that—within the margins of uncertainty—neither diuretic confers a clearly superior survival advantage in any ISTE-defined patient stratum.

**Figure 3.**
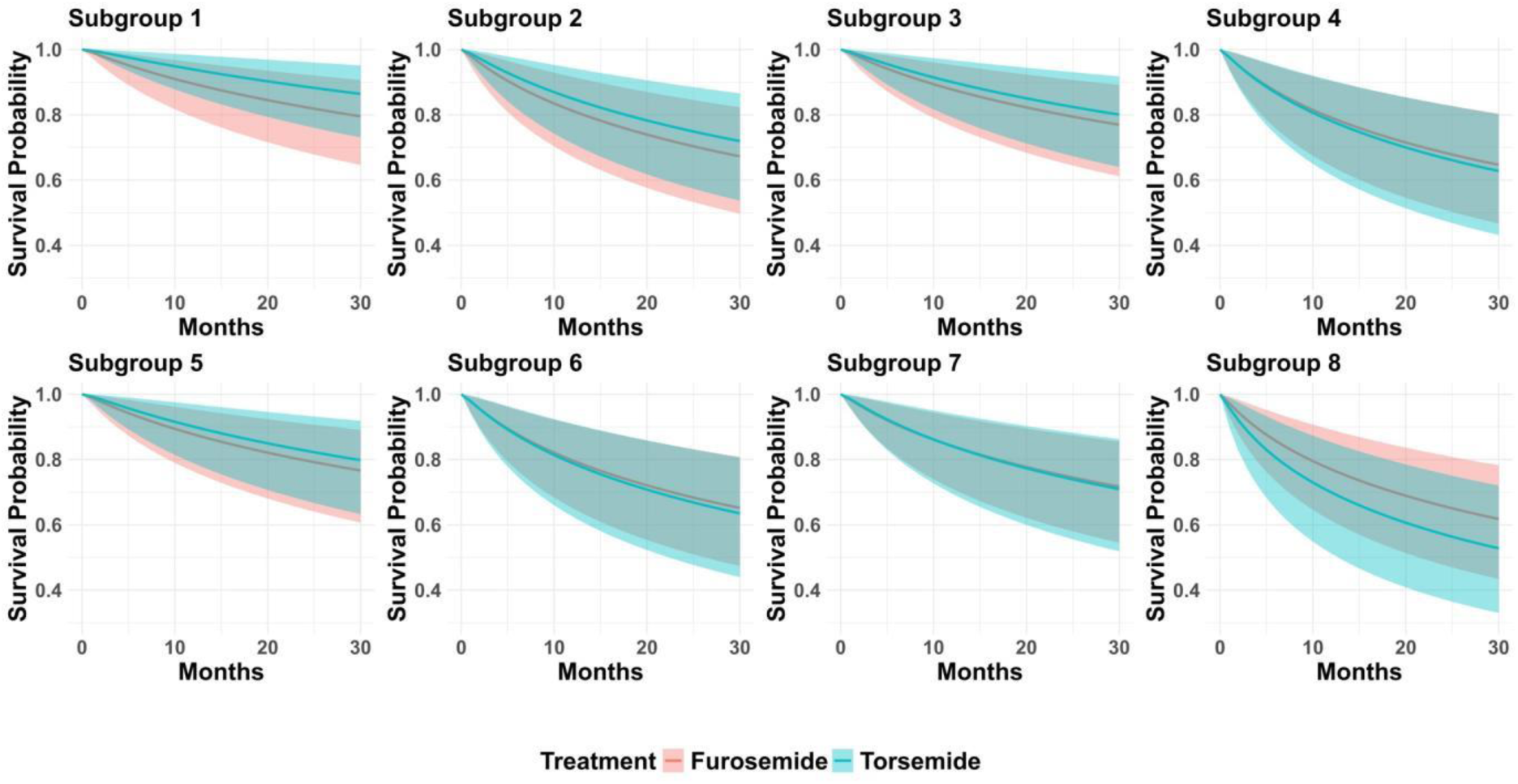
Survival probability curves under torsemide versus furosemide in each group. Note: Subgroups correspond to the eight groups in Figure 2. **Subgroup 1**: without atrial fibrillation/flutter, std. BNP < −0.25, no prior loop diuretic prescription; **Subgroup 2**: without atrial fibrillation/flutter, std. BNP < −0.25, prior loop diuretic prescription; **Subgroup 3**: without atrial fibrillation/flutter, std. BNP ≥ −0.25, no prior loop diuretic prescription; **Subgroup 4**: without atrial fibrillation/flutter, std. BNP ≥ −0.25, prior loop diuretic prescription; **Subgroup 5**: with atrial fibrillation/flutter, std. BNP < −0.25, no prior loop diuretic prescription; **Subgroup 6**: with atrial fibrillation/flutter, std. BNP < −0.25, prior loop diuretic prescription; **Subgroup 7**: with atrial fibrillation/flutter, std. BNP ≥ −0.25, no prior loop diuretic prescription; **Subgroup 8**: with atrial fibrillation/flutter, std. BNP ≥ −0.25, prior loop diuretic prescription.

**Table 3** reports the cumulative hazard ratios (CHRs) for torsemide versus furosemide in each of the eight subgroups defined by individual survival treatment effects (ISTE), evaluated at both 12 months and 30 months. We focus on CHRs rather than conventional hazard ratios because CHRs admit a valid causal interpretation even when the proportional-hazards assumption does not hold within subgroups. Although the point estimates vary—ranging from modest reductions in hazard (e.g. CHR≈0.78 in the subgroup most favoring furosemide) to slight elevations (e.g. CHR≈1.76 in the subgroup most favoring torsemide) at one year—the 95% credible intervals for every subgroup at both time horizons include 1.0. This consistent overlap with the null value indicates that, despite suggestive heterogeneity in the estimated effects across patient profiles, there is no statistically or credibly significant difference in cumulative mortality hazard between torsemide and furosemide in any subgroup.

**Table 3.** Cumulative hazard ratios for subgroups at 12 and 30 months.

|  | Subgroups |  |  |  |  |  |  |  |
| --- | --- | --- | --- | --- | --- | --- | --- | --- |
|  | 1 | 2 | 3 | 4 | 5 | 6 | 7 | 8 |
| No. treated/total | 144/290 | 212/428 | 174/338 | 276/529 | 77/171 | 224/465 | 72/140 | 252/498 |
| **12 Months** |  |  |  |  |  |  |  |  |
| Mortality in Furosemide | 15/146 | 34/216 | 20/164 | 49/253 | 12/94 | 36/241 | 10/68 | 61/246 |
| Mortality in Torsemide | 6/144 | 23/212 | 12/174 | 68/276 | 5/77 | 37/224 | 11/72 | 79/252 |
| CHR [95% credible interval] | 0.78[0.12, 2.66] | 0.99[0.23, 2.88] | 1.11[0.19, 3.65] | 1.4[0.34, 3.92] | 1.12[0.2, 3.65] | 1.35[0.34, 3.68] | 1.44[0.3, 4.41] | 1.76[0.49, 4.63] |
| **30 Months** |  |  |  |  |  |  |  |  |
| Mortality in Furosemide | 20/146 | 53/216 | 26/164 | 77/253 | 16/94 | 64/241 | 16/68 | 86/246 |
| Mortality in Torsemide | 11/144 | 36/212 | 23/174 | 93/276 | 11/77 | 63/224 | 15/72 | 107/252 |
| CHR [95% credible interval] | 0.79[0.17, 2.3] | 0.98[0.3, 2.46] | 1.07[0.26, 3.01] | 1.31[0.41, 3.18] | 1.07[0.26, 3] | 1.27[0.42, 3] | 1.33[0.37, 3.52] | 1.6[0.56, 3.68] |

**Note:** Subgroups correspond to the eight groups in Figure 2. **Subgroup 1:** without atrial fibrillation/flutter, std. BNP < -0.25, no prior loop diuretic prescription; **Subgroup 2:** without atrial fibrillation/flutter, std. BNP < -0.25, prior loop diuretic prescription; **Subgroup 3:** without atrial fibrillation/flutter, std. BNP ≥ -0.25, no prior loop diuretic prescription; **Subgroup 4:** without atrial fibrillation/flutter, std. BNP ≥ -0.25, prior loop diuretic prescription; **Subgroup 5:** with atrial fibrillation/flutter, std. BNP < -0.25, no prior loop diuretic prescription; **Subgroup 6:** with atrial fibrillation/flutter, std. BNP < -0.25, prior loop diuretic prescription; **Subgroup 7:** with atrial fibrillation/flutter, std. BNP ≥ -0.25, no prior loop diuretic prescription; **Subgroup 8:** with atrial fibrillation/flutter, std. BNP ≥ -0.25, prior loop diuretic prescription.

Overall, torsemide appears most beneficial among patients with no history of AF, lower BNP/NTproBNP, and no prior loop diuretic prescription (Subgroup 1 treatment effect = 0.49, 95% CrI= [-0.07, 1.04]), while furosemide is more favored among patients with atrial fibrillation/flutter, higher BNP/NTproBNP, and prior loop diuretic prescription (Subgroup 8 treatment effect = -0.49, 95% CrI= [-0.86, -0.13]). Using this decision framework in TRANSFORM-HF patients was associated with a model-estimated improvement in median survival among deceased patients from 7.9 to 9.6 months compared to randomization, corresponding to a relative increase of about 22%. Compared with a “torsemide for all” strategy, this corresponds to a 24.3% extension, and compared with a “furosemide for all” strategy, a 20.7% extension. While these projections are exploratory, hypothesis-generating, and likely overstate the magnitude of any true clinical effect, they suggest that individualized treatment assignment could potentially offer greater benefit than a uniform “torsemide for all” or “furosemide for all” strategy.

## Discussion

This study applied a machine learning framework to illustrate the HTE detection in TRANSFORM-HF trial. Using a Bayesian AFT-BART framework, we incorporated over 50 baseline patient characteristics and observed meaningful heterogeneity in treatment effects between torsemide and furosemide in TRANSFORM-HF. The exploratory decision tree identified AF, BNP/NT-proBNP levels, and prior loop diuretic use as the most influential baseline modifiers of survival benefit. These findings suggest that patient phenotype may influence diuretic response more than has been appreciated in prior analyses.

Our results align with pathophysiological findings demonstrated in the TRANSFORM-HF mechanistic substudy.^20^ Patients with atrial fibrillation and elevated natriuretic peptides often present with greater congestion and altered renal hemodynamics, settings in which furosemide’s pharmacodynamic profile, including longer renal drug delivery and more sustained natriuresis, may confer a relative advantage. In contrast, while torsemide is frequently cited for more predictable oral absorption and longer half-life (e.g., ^12^), the same mechanistic study showed that higher torsemide dosing was associated with mild renal impairment and marked neurohormonal activation (renin, aldosterone, norepinephrine).^20^ Thus, the theoretical pharmacokinetic advantages of torsemide did not uniformly translate into more favorable decongestion in real-world heart failure patients. Although torsemide has more predictable oral absorption and a longer elimination half-life in healthy volunteers, higher torsemide dosing in contemporary patients was actually associated with mild perturbations in renal function and significant increases in renin, aldosterone, and norepinephrine. Therefore, despite its bioavailability, torsemide does not uniformly translate into a “smoother” diuresis in real-world heart failure patients.

Among the three factors identified in the explanatory decision tree, AF was the dominant splitter: patients without AF tended to fare better on torsemide, whereas those with AF trended toward better survival on furosemide. AF is consistently linked to greater congestion, renal vulnerability, and worse outcomes in HF patients,^21^ a phenotype in which a stronger natriuretic profile may be advantageous. Within each AF stratum, higher BNP/NT-proBNP further shifted the ISTE toward furosemide, perhaps because natriuretic peptides track congestion severity and strongly predict adverse outcomes, this gradient is biologically plausible.^22,23^ Mechanistically, the TRANSFORM-HF mechanistic substudy reported longer renal drug delivery and more sustained natriuresis with furosemide than with torsemide—especially relevant in more congested states—while higher torsemide exposure was associated with greater neurohormonal activation (renin, aldosterone, norepinephrine).^20^ A third modifier was prior loop-diuretic use, where non-users showed suggestive benefit with torsemide, but prior users tended to have attenuated effects. Prior exposure likely indexes dose intensity and diuretic resistance, and recent post-hoc TRANSFORM-HF dose analyses show that higher loop-diuretic doses—regardless of agent—track with worse outcomes, underscoring that dose and disease severity can confound agent comparisons.^24^

Taken together, our findings suggest that patients with atrial fibrillation and higher BNP levels appear to derive relatively greater benefit from furosemide, whereas those without AF, with lower BNP levels, and without prior loop diuretic exposure may have more favorable responses to torsemide. This pattern is biologically plausible, as the TRANSFORM-HF mechanistic substudy demonstrated that furosemide provided more sustained renal drug delivery and natriuresis, whereas higher torsemide dosing was associated with neurohormonal activation and mild renal impairment—features that may be particularly consequential in patients with more severe congestion.^20^ In addition, AF and elevated natriuretic peptides often reflect more severe volume overload and altered renal hemodynamics, settings in which furosemide’s pharmacodynamic profile may confer an advantage.^25^ Finally, because TRANSFORM-HF was explicitly designed as a pragmatic randomized trial, our findings underscore why pragmatic cardiovascular trials should plan for and report heterogeneity of treatment effects (HTE) rather than focus solely on a single average treatment effect. Pragmatic trials aim to estimate real-world effectiveness under usual care—using broad eligibility, flexible intervention delivery, and patient-centered outcomes—thereby maximizing applicability but also introducing multiple sources of heterogeneity across patients and settings.^29,30^ Contemporary guidance (e.g., CONSORT-Pragmatic and PRECIS-2) emphasizes applicability and transparent reporting as well as recommends pre-specifying the role of HTE analyses (both confirmatory and exploratory) and interpreting results with attention to multiplicity and transportability.^30^ Reviews focused on pragmatic trials highlight that heterogeneity is intrinsic in these designs and should be anticipated and managed analytically; indeed, pragmatic trials are increasingly used in patient-centered outcomes research specifically to assess HTE.^27^ Our standardized workflow for HTE in pragmatic trials—demonstrated here in TRANSFORM-HF—seeks to operationalize these principles by promoting transparent pre-specification, robust estimation, and clinically interpretable summaries that can inform future confirmatory studies and practice.

## Limitations

While our work is intended to showcase the HTE detection in pragmatic cardiovascular trials, several limitations in our illustration with TRANSFORM HF are worth nothing. To note, because the torsemide arm received approximately twice the total diuretic dose, it remains unclear whether observed differences reflect dose effects, drug-specific effects, or both. A recent post-hoc analysis of TRANSFORM-HF has shown that higher loop diuretic doses—whether torsemide or furosemide—were consistently associated with worse outcomes, including increased mortality and hospitalization, without evidence of heterogeneity by drug type.^24^ Thus, further investigation is needed before concluding that loop diuretics are not interchangeable or to guide personalized diuretic selection. Another limitation of this work is that we have only applied our machine learning approach to a single trial, but further validation in external datasets is necessary to refine precision medicine approaches in diuretic therapy. Relatedly, the TRANSFORM HF findings are exploratory and hypothesis-generating, not intended to inform clinical guidelines at this stage. Future trials and studies using independent data sources should evaluate treatment effect heterogeneity between torsemide and furosemide in identified strata and determine whether differential prescribing of these agents may be warranted.

## Data Availability

Data is available through request at Biolincc.

## Abbreviations

ACE: Angiotensin-Converting Enzyme
AFT: Accelerated Failure Time
AI: Artificial Intelligence
AF: Atrial Fibrillation
AFIBN: Atrial Fibrillation/Flutter
ARNi: Angiotensin Receptor-Neprilysin Inhibitor
BART: Bayesian Additive Regression Tree
BMI: Body Mass Index
BNP: B-type Natriuretic Peptide
CART: Classification and Regression Tree
CATE: Conditional Average Treatment Effect
CI: Confidence Interval
CrI: Credible Interval
GDMT: Guideline-Directed Medical Therapy
GLP-1: Glucagon-Like Peptide-1
HF: Heart Failure
HFrEF: Heart Failure with Reduced Ejection Fraction
HFmrEF: Heart Failure with Mildly Reduced Ejection Fraction
HFpEF: Heart Failure with Preserved Ejection Fraction
HR: Hazard Ratio
ISTE: Individual Survival Treatment Effect
KCCQ: Kansas City Cardiomyopathy Questionnaire
LDPRIORN: Prior Loop Diuretic Use
LVEF: Left Ventricular Ejection Fraction
MCMC: Markov Chain Monte Carlo
MRA: Mineralocorticoid Receptor Antagonist
NT-proBNP: N-terminal Prohormone of B-type Natriuretic Peptide
NYHA: New York Heart Association
RR: Rate Ratio
SGLT2: Sodium-Glucose Cotransporter-2
TRANSFORM-HF: Torsemide Comparison with Furosemide for Management of Heart Failure

## Acknowledgement

We thank participants, staff, and funders of TRANSFORM HF.

